# Iron metabolism disorders of patients with chronic paracoccidioidomycosis

**DOI:** 10.1101/2023.02.10.23285778

**Authors:** Eliana da Costa Alvarenga de Brito, Igor Valadares Siqueira, James Venturini, Vinícius Lopes Teodoro Félix, Alana Oswaldina Gavioli Meira dos Santos, Rinaldo Poncio Mendes, Simone Schneider Weber, Anamaria Mello Miranda Paniago

## Abstract

Paracoccidioidomycosis (PCM) is caused by *Paracoccidioides* spp.; during infection, some host mechanisms limit the availability of iron, thereby reducing its reproduction. However, *Paracoccidioides* spp. can evade the immune defense and, even under limited iron conditions, use this mineral for growth and dissemination. This study evaluated the iron metabolism of 39 patients who were diagnosed with chronic PCM between 2013 and 2021. The forms of iron before treatment and at the time of clinical cure were evaluated based on the following: serum ferritin levels (storage iron); total iron-binding capacity (TIBC) and transferrin saturation (TSAT) level (transport iron); red blood cell (RBC), hemoglobin (Hb), hematocrit (HCT), and soluble transferrin receptor (sTfR) levels; and sTfR/log ferritin ratio (functional iron). The mean age of the patients was 54.5 years (±6.7 years). Most patients were men (97.4%), rural workers (92.1%), and smokers (84.6%); most had moderate disease severity (66.7%). Before treatment, the median values of all evaluated parameters were within or just slightly outside the normal range of values. However, it is possible to infer that PCM interferes with functional and storage iron because improvements in these parameters after treatment as well as associations with disease severity were observed. Furthermore, moderate correlations were observed between C-reactive protein and the Hb (r=-0.500; p=0.002), RBC (r=-0.461; p=0.005), HCT (r=-0.514; p=0.001), and iron levels (r=-0.491; p=0.002). PCM interferes with iron metabolism by transforming functional iron to storage iron, as revealed by anemia, low iron levels with normal TSAT levels, normal TIBC, normal sTfR levels, normal sTfR/log ferritin ratios, and normal or slightly increased ferritin levels. PCM can lead to anemia of inflammation, which can be differentiated from iron deficiency anemia by a careful investigation of the iron form parameters.

## Introduction

Paracoccidioidomycosis (PCM), which is a systemic mycosis endemic to Latin America, is caused by dimorphic fungi, *Paracoccidioides* spp. Infection occurs through the inhalation of infective conidia that are present in the soil; these conidia settle in the lungs and can spread to the lymphatic and hematogenous pathways and other organs. Brazil is the main endemic area for PCM, which is considered the eighth leading cause of mortality associated with chronic infectious diseases in the country [1]. Chronic or adult PCM, which is observed in those older than age 30 years, is the most frequent form of this disease, and it predominantly affects the lungs and mucous membranes of the upper digestive tract and airways [2].

Similar to other fungal diseases, certain factors are determinants of PCM, such as the inoculum size, pathogenicity, fungus virulence, defense system integrity, and genetic factors [3]. The host organism uses strategies or barriers that prevent fungal diseases, such as inhibiting vital fungal functions by blocking iron uptake; this is because iron is an essential mineral for the proliferation of many pathogens, including *Paracoccidioides* spp. [4].

Iron is an essential component of hemoglobin (Hb), myoglobin, and several enzymes, and it has a fundamental role in oxygen transport and electron transfer. Additionally, it acts as a cofactor in many enzymatic processes, including DNA synthesis [5-7].

Maintaining control of iron homeostasis is essential in host-pathogen interactions because both compete for this micronutrient. During infection, some host immune mechanisms limit the availability of iron to invading microorganisms, thereby reducing their proliferation [8]. However, many microorganisms, such as *Paracoccidioides* spp., can evade the immune defense; even under limited iron conditions, they can use this mineral for growth and dissemination [9]. Therefore, alterations in hematological parameters and iron metabolism can be observed during PCM infection [10].

In the human body, iron is distributed in different interconnected forms to enable homeostasis. These forms of iron (storage iron, transport iron, and functional iron) can be evaluated separately using laboratory tests, many of which are available and accessible during routine laboratory testing. These tests can assess any disturbances in iron metabolism and the affected iron form [11]. Generally, when there is an iron deficit, these forms are sequentially affected. First, there is a decrease in the storage iron levels, followed by a deficiency in the transport iron level and a reduction in the functional iron level [12,13].

The host-parasite interaction and consequent inflammatory response lead to changes in iron metabolism dynamics. These changes can be observed in some patients with PCM [14], which is a chronic systemic inflammatory disease requiring long-term treatment. Understanding the frequency and intensity of iron metabolism disturbances in patients with chronic PCM will help improve therapeutic management. Therefore, we analyzed the iron metabolism and different iron forms of patients with chronic PCM before treatment and at the time of clinical cure.

## Materials and Methods

### Ethical aspects

This study was approved by the Human Research Ethics Committee of the Federal University of Mato Grosso do Sul (CAAE number 62726016.2.0000.0021). All participants provided written informed consent.

### Location, period, and design of the study

This study was performed between 2013 and 2021 at the outpatient clinic for systemic mycoses of the Infectious and Parasitic Diseases Unit of the University Hospital Maria Aparecida Pedrossian at the Federal University of Mato Grosso do Sul, which is a reference center for infectious diseases, in Campo Grande, Mato Grosso do Sul, Brazil. This analytical study involving prospective data evaluated iron metabolism parameters of patients with chronic PCM before treatment and at the time of clinical cure.

### Patients

#### Inclusion and exclusion criteria

Male and female individuals who were diagnosed with chronic PCM between September 2013 and February 2021 were included. Patients with chronic PCM who died and/or did not undergo laboratory tests at admission, such as serum iron and blood count (hemogram [HMG] test) tests, were excluded, as were patients diagnosed with human immunodeficiency virus, tuberculosis, and/or neoplasia.

#### Case definition

Confirmed cases of PCM were those presenting with suggestive clinical manifestations, typical yeast forms of *Paracoccidioides* spp. visualized by mycological, culture, or direct histopathological tests, and/or serum-specific antibodies detected using the double agar gel immunodiffusion test.

### Demographic and clinical variables

Demographic, clinical, and laboratory data were retrieved from a prospective database. They comprised complete routine medical data obtained at the time of diagnosis and during follow-up of the disease. Demographic data, such as sex, age, occupational history, smoking, and clinical disease severity, were collected before treatment.

Chronic PCM was subdivided into mild, moderate, and severe cases. Mild cases included body mass index (BMI) reduction <5% of the usual BMI and the involvement of one or a few organs without functional changes. Severe cases comprised three or more of the following criteria: BMI reduction ≥10% of the usual BMI; severe pulmonary impairment; involvement of other organs, such as the adrenal glands, central nervous system, and bones; enlargement of multiple lymph node chains (superficial or deep chains with pseudotumors [>2.0 cm in diameter] with or without suppuration); and high-titer antibodies. Moderate cases were defined as intermediate cases that were worse than mild cases but not as serious as severe cases [2].

### Laboratory analysis

Blood samples were obtained during the routine medical examination at the time of diagnosis and during follow-up. Laboratory data and complementary laboratory test data were obtained at two stages of routine outpatient follow-up: before treatment and at the time of clinical cure (disappearance of signs and symptoms) [2]. To investigate the iron metabolism of patients with chronic PCM, different forms of iron were evaluated.

#### Storage iron

Storage iron is iron stored in the body. Serum ferritin levels were analyzed using an electrochemiluminescence immunoassay (Roche).

#### Transport iron

Transport iron is iron that circulates in the body. The total iron-binding capacity (TIBC) was measured using a colorimetric assay (Roche) and the transferrin saturation (TSAT) level, which was calculated as [TSAT = serum iron/TIBC × (100)]; the result was expressed as a percentage (%).

#### Functional iron

Functional iron is related to erythrocyte synthesis and erythropoiesis. Therefore, to evaluate functional iron, the blood count (HMG test) was evaluated to measure the Hb level, red blood cell (RBC), level mean corpuscular volume, and mean Hb concentration (MHC) (XN 3000 series hematology equipment; Sysmex Corporation, Kobe, Japan). Serum iron was determined using a colorimetric assay (Roche). The soluble transferrin receptor (sTfR) level was determined using an enzyme-linked immunosorbent assay (human sTfR ELISA Kit; Elabscience). This test is not routinely performed; therefore, serum stored in the PCM biobank was used.

The inflammatory process was assessed according to the high-sensitivity serum C-reactive protein (CRP) level measured using immunoturbidimetry. All analyzed parameters and values considered normal by the manufacturer are listed in **Table 1**.

**Table 1.**
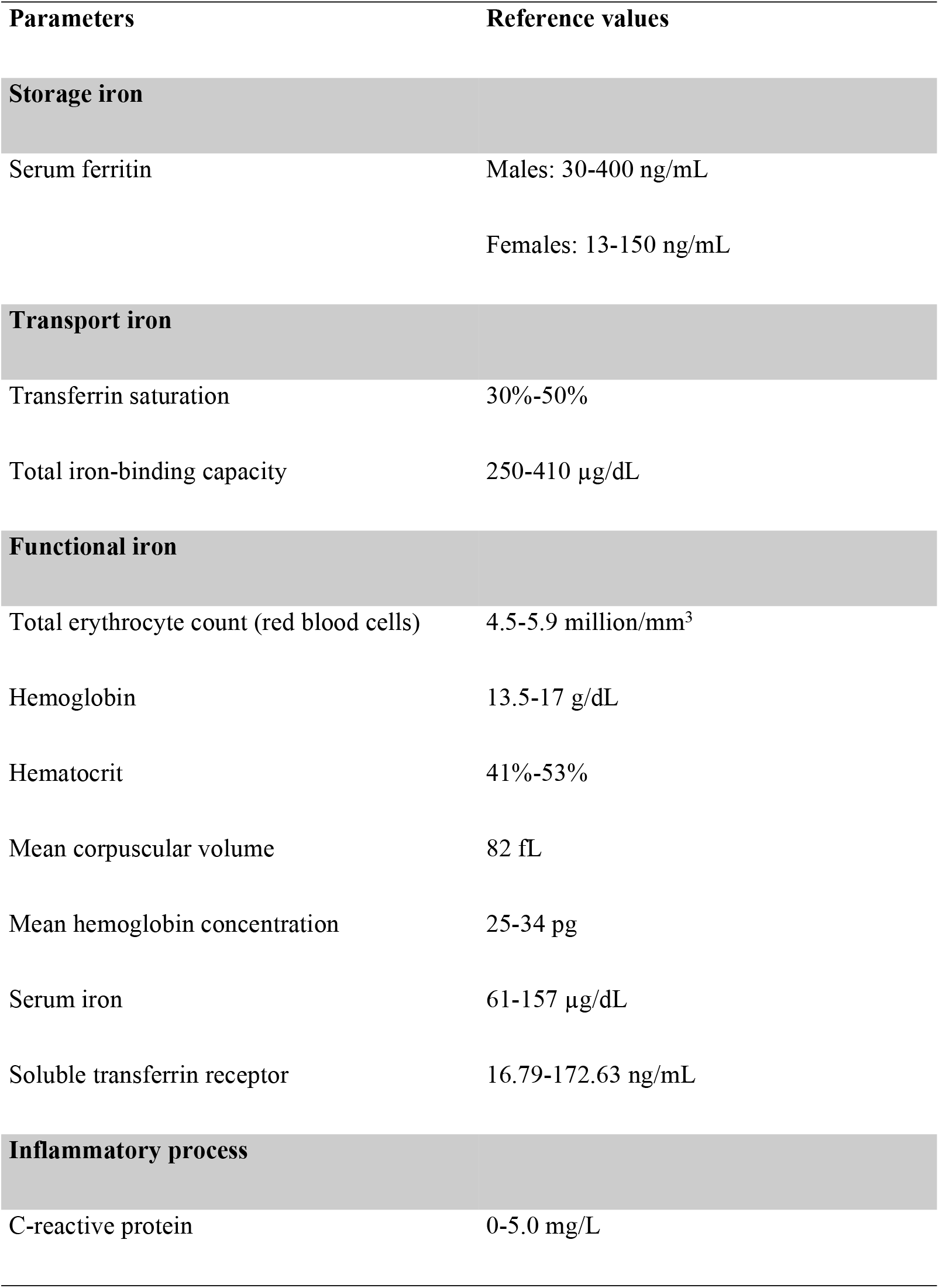
Iron parameters and reference values provided by the manufacturer.

### Statistical analysis

The statistical data analysis was performed using Jamovi software (version 1.6) for Windows [15]. Data are presented as the mean ± standard deviation (SD) or median with the first quartile (Q1) and third quartile (Q3). Comparisons between values before treatment and at the time of clinical cure were performed using Student’s t test or the Wilcoxon signed rank test. The Pearson correlation coefficient I was used to determine the relationship between variables; values ≤0.29, between 0.30 and 0.49, and between 0.50 and 1.0 were considered weak, moderate, and strong correlations, respectively.

McNemar’s test was performed to assess the associations between qualitative variables. The Shapiro-Wilk test was performed to determine whether the variables were normally distributed.

A proportion test and one-sample test were performed to analyze the categorical and numerical variables, respectively. For all tests, p ≤ 0.05 was considered significant.

## Results

A total of 88 patients were diagnosed with chronic PCM during the study period. Of these, 35 patients were excluded according to the criteria and 39 patients participated in the study (**Fig 1**). The mean age of these patients was 54.5 years (SD, 6.70 years); 38 (97.4%) were men and 35 (92.1%) were rural workers. Thirty-three (84.6%) individuals were smokers at the time of diagnosis. Regarding disease severity, 66.7% of the patients had moderate disease, 20.5% had severe disease, and 12.8% had mild disease (**Table 2**).

**Fig 1.**
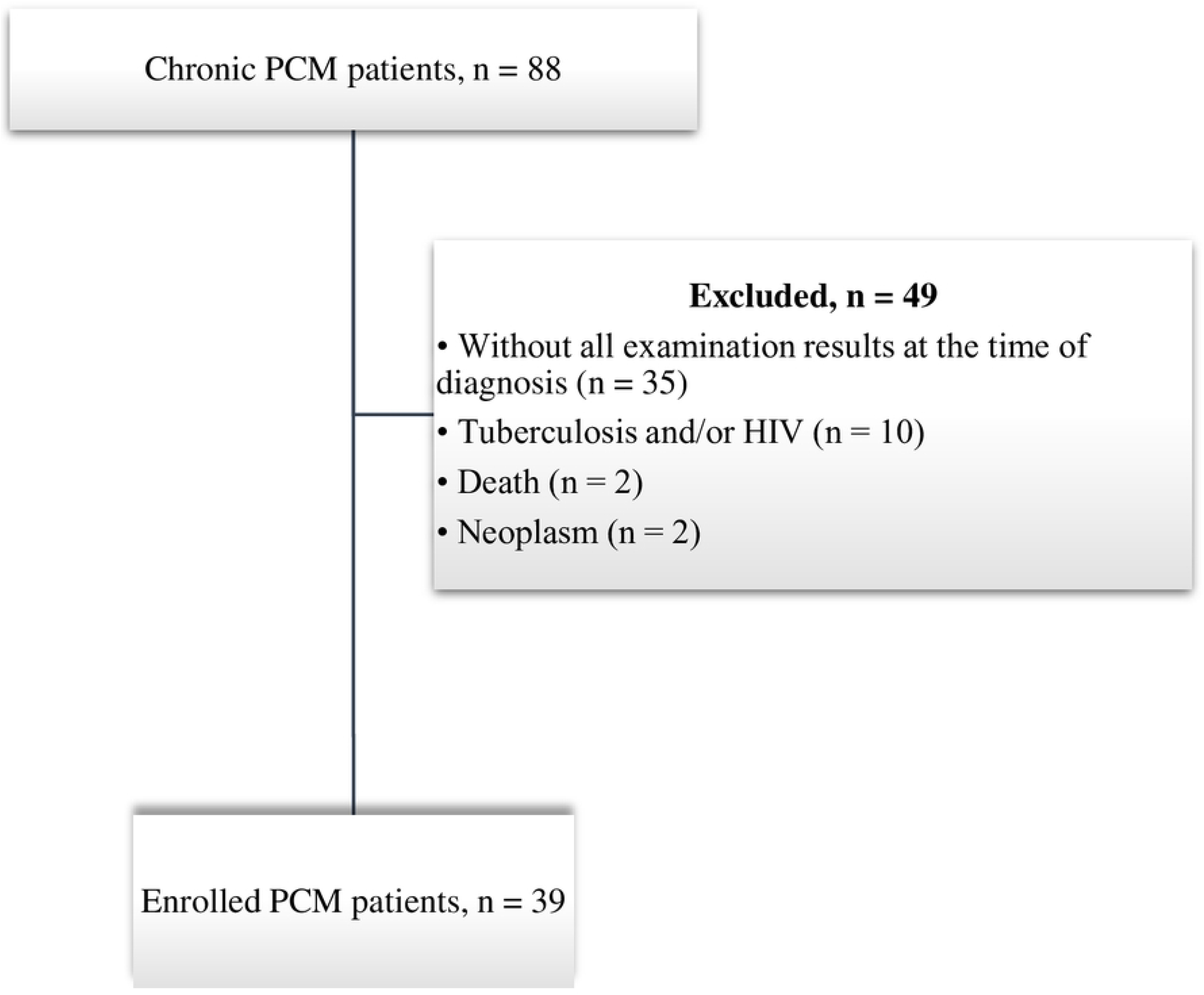
Flowchart of the selection of participants with chronic paracoccidioidomycosis (PCM) from 2013 to 2021. HIV, human immunodeficiency virus.

**Table 2.**
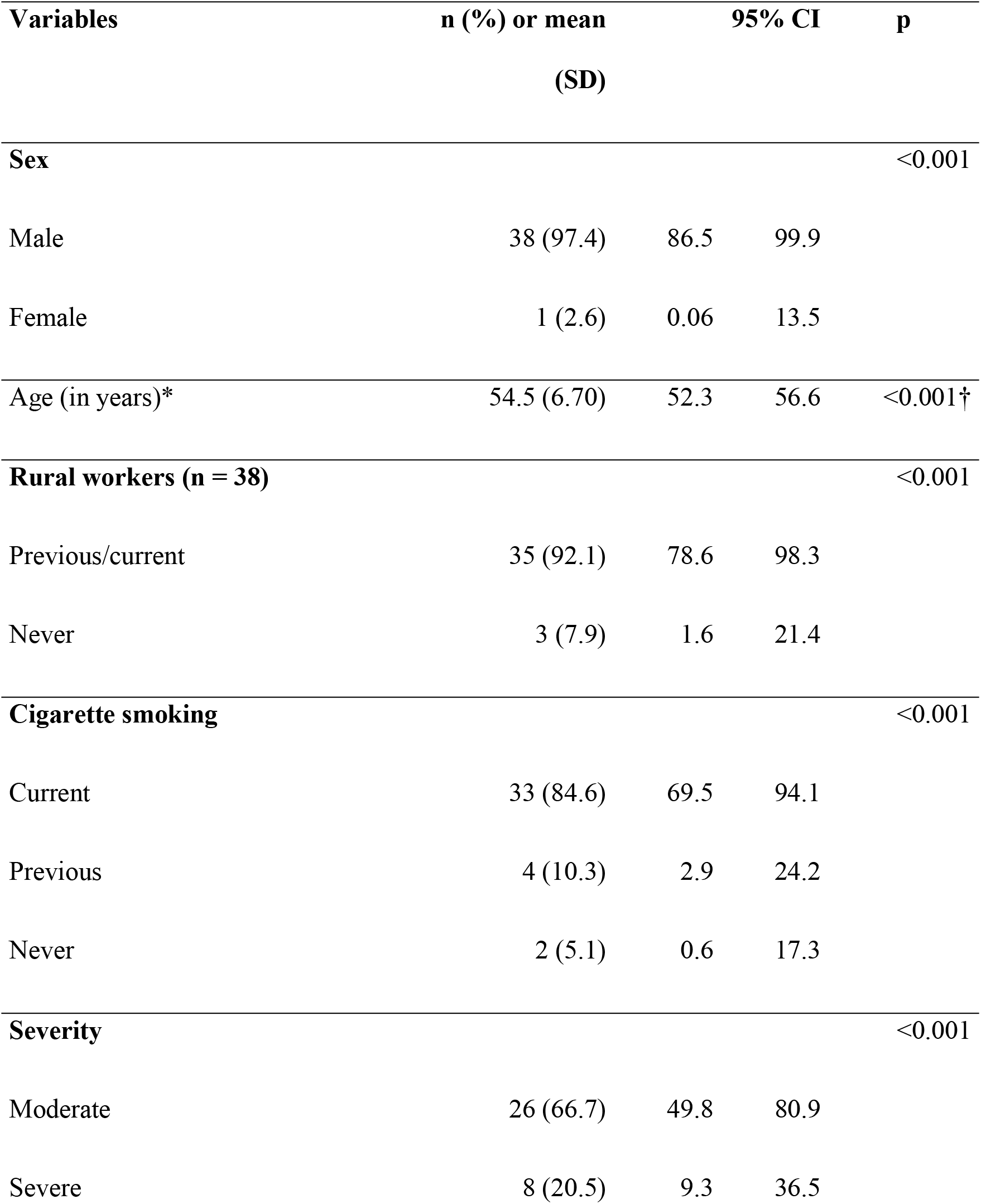
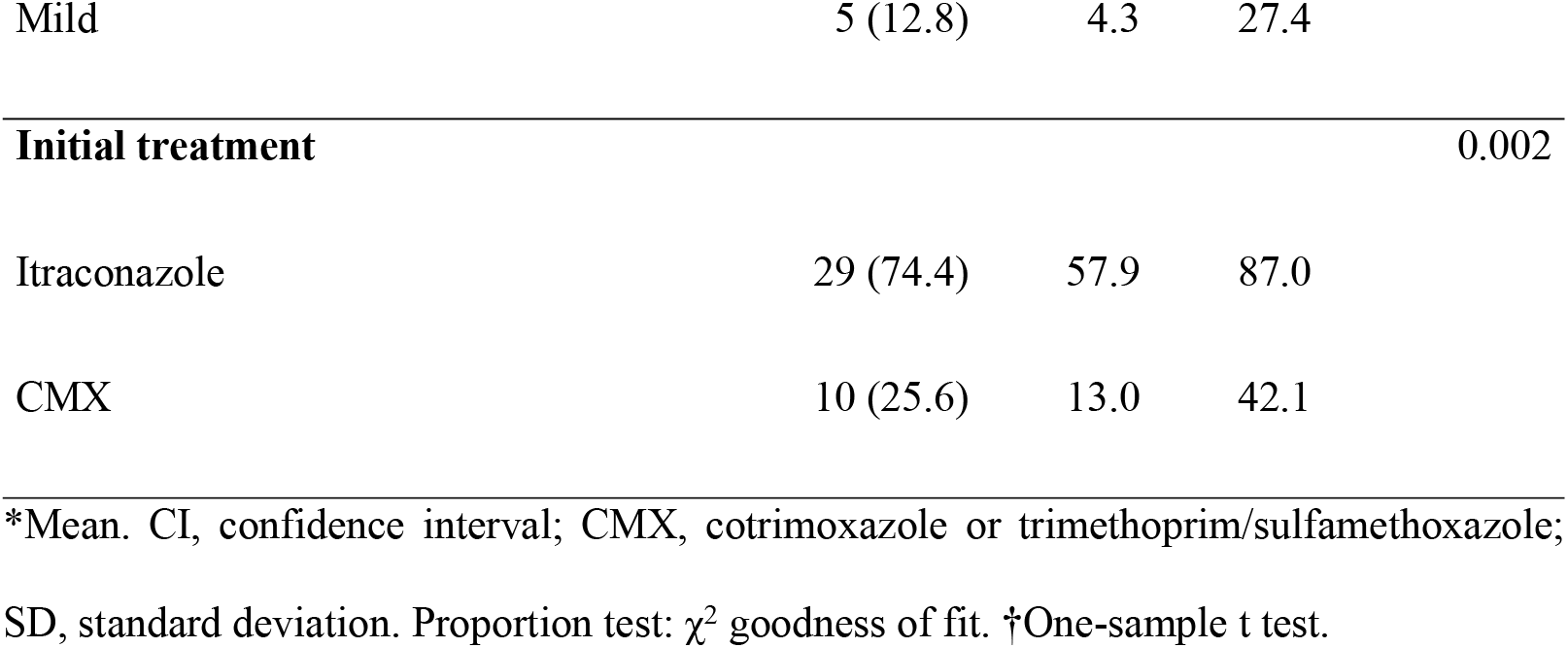
Baseline demographic, clinical, and lifestyle data of 39 patients with chronic paracoccidioidomycosis.

### Storage iron

Nine (26.5%) patients had increased ferritin levels and two (5.8%) patients had lower than normal ferritin levels before treatment (evaluated before treatment: n = 34 patients). An analysis of patients who were evaluated at both time points (before treatment and at the time of clinical cure: n = 28 patients) showed that the number of individuals with high ferritin levels decreased (21.4% before treatment vs. 17.9% at the time of clinical cure; p = 0.008). An analysis of the median values showed that those at the time of clinical cure were reduced compared to those before treatment (p = 0.017) (**Fig 2**).

**Fig 2.**
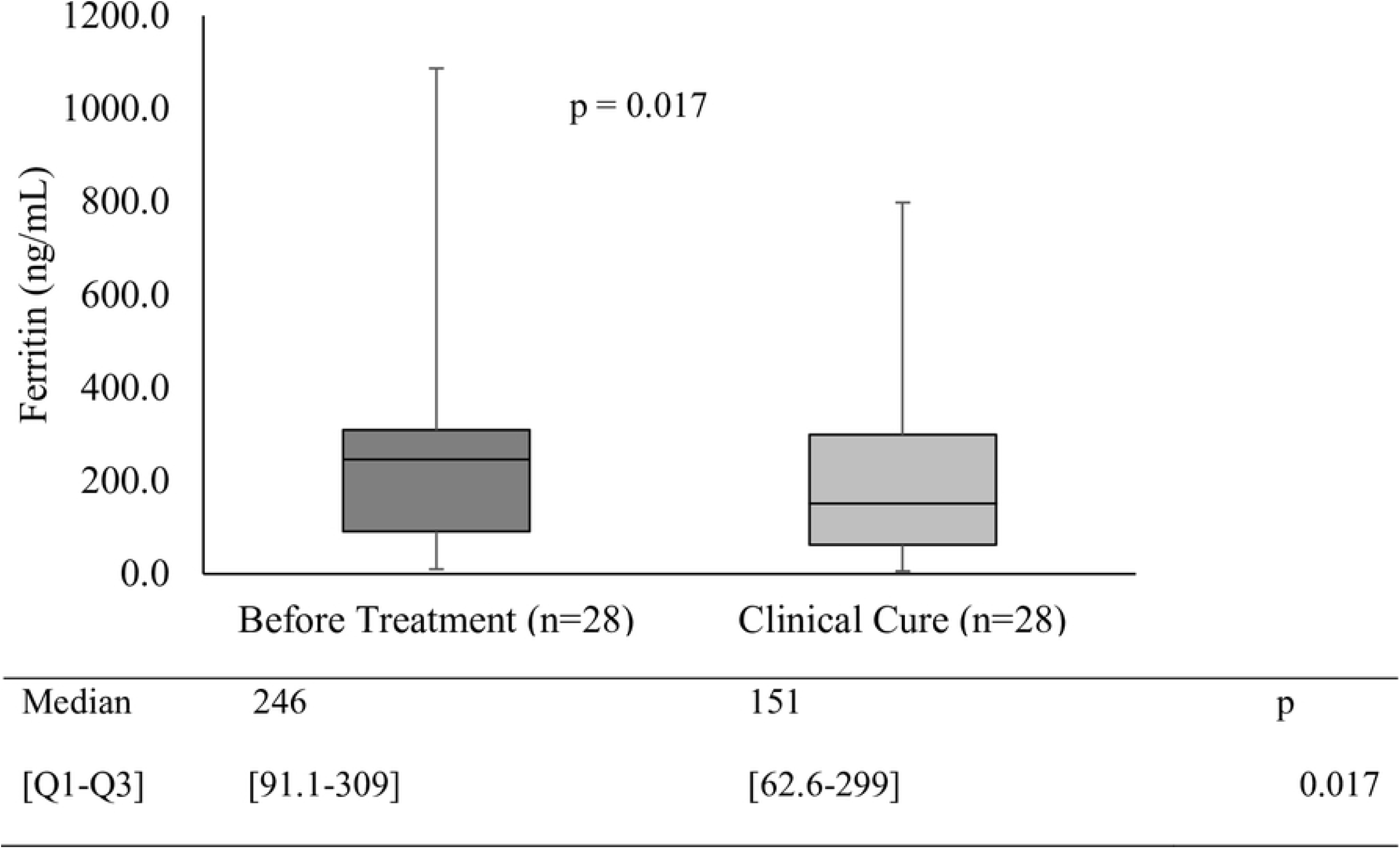
Comparison of median serum ferritin values before treatment (BT) and at the time of clinical cure (CC) of patients with chronic paracoccidioidomycosis. Q1, first quartile; Q3, third quartile. Wilcoxon signed rank test.

### Transport iron

Transport iron was evaluated according to the TSAT level and TIBC. Of the 39 patients evaluated before treatment, 27 (69.2%) had TSAT levels <30% and 8 (20.5%) had TSAT levels <16%. At the time of clinical cure, the number of patients with TSAT levels <30% (n = 12; 42.9%) did not decrease (before treatment vs. clinical cure: n = 28; p = 0.071). Similarly, an analysis of the median TSAT values showed no statistical differences between those before treatment and those at the time of clinical cure (**Table 3**).

**Table 3.** Comparison of mean or median values of the transport iron parameters before treatment and at the time of clinical cure of 30 patients with chronic paracoccidioidomycosis.

| Variables | Before Treatment |  | Clinical Cure |  | p |
| --- | --- | --- | --- | --- | --- |
|  | Mean or | SD or | Mean or | SD or |  |
|  | Median | Q1-Q3 | Median | Q1-Q3 |  |
| TSAT (%) | 26 | 18.0 -36.8 | 32.5 | 24.3-40.0 | 0.076 |
| TIBC (µg/dL) | 245 | 100.9 | 265 | 85.7 | 0.398 |

Q1, first quartile; Q3, third quartile; SD, standard deviation; TIBC, total iron-binding capacity; TSAT, transferrin saturation. The Wilcoxon signed rank test and paired Student's t test were performed.

The TIBC analysis showed that 22 (56.4%) individuals had lower than normal values before treatment (n = 39). At the time of clinical cure, the number of individuals with TIBC lower than the reference value had not decreased (n = 10; 35.7%; p = 0.439). An analysis of the mean values before treatment showed that the TIBC was slightly lower than normal, and that it had not increased at the time of clinical cure (**Table 3**).

### Functional iron

The HMG test was performed to evaluate the Hb, RBC, and HCT levels, mean corpuscular volume, and mean corpuscular Hb level to assess functional iron. Additionally, to complement this evaluation, the serum iron and sTfR values and sTfR/log ferritin ratio were calculated.

The HMG test results before treatment (n = 39) showed that 36% (n = 14) of individuals had reduced Hb, RBC, and HCT levels. Anemia was considered when the Hb level was <13.5 g/dL. It was noted that 46.1% (n = 18) of patients had anemia; among these patients, 72% (n = 13) had normocytic and normochromic anemia. The mean Hb level among the 18 patients with anemia was 11.2 g/dL (SD, 1.21 g/dL).

During the healing process of these patients, the anemia frequency and HCT levels were significantly reduced (**Table 4**). These improvements with treatment were also observed when evaluating the mean values, which were significantly increased at the time of clinical cure compared to those before treatment (**Table 5**).

**Table 4.** Changes in functional iron parameters at the time of clinical cure compared to those before treatment of 35 patients with chronic paracoccidioidomycosis.

| Variables | Before Treatment | Clinical Cure | p |
| --- | --- | --- | --- |
|  | n (%) | n (%) |  |
| Anemia | 14 (40.0) | 6 (17.1) | 0.011 |
| Reduced RBCs | 16 (45.7) | 11 (31.4) | 0.132 |
| Reduced HCT | 18 (51.4) | 11 (31.4) | 0.035 |
| Reduced iron* | 15 (51.7) | 4 (12.5) | <0.001 |

\*Iron evaluation of 32 patients. The McNemar test was performed. HMG: hemogram; RBCs, red blood cells (total erythrocyte count); HCT, hematocrit.

**Table 5.** Comparison of functional iron parameter values before treatment and at the time of clinical cure of patients with chronic paracoccidioidomycosis.

| Variables | Before Treatment |  |  | Clinical Cure |  |  | p |
| --- | --- | --- | --- | --- | --- | --- | --- |
|  | n | Mean or<br>Median | SD or<br>Q1-Q3 | n | Mean or<br>Median | SD or<br>Q1-Q3 |  |
| Hb | 35 | 13.1 | 2.2 | 35 | 14.5 | 1.8 | <0.001 |
| RBCs | 35 | 4.5 | 0.6 | 35 | 4.8 | 0.6 | 0.002 |
| HCT | 35 | 39.9 | 6.1 | 35 | 43.6 | 5.2 | <0.001 |
| MCV | 35 | 89.0 | 5.7 | 35 | 91.0 | 5.3 | 0.093 |
| MCH | 35 | 29.3 | 2.3 | 35 | 30.4 | 2.2 | 0.008 |
| Iron | 32 | 64.0 | 37.0-88.5 | 32 | 85.5 | 69.0-109.0 | 0.004 |
| sTfR | 18 | 21.5 | 17.4-7.6 | 18 | 16.2 | 6.3-20.2 | 0.099 |
| sTfR/logFerr | 11 | 8.1 | 2.8 | 11 | 7.6 | 3.9 | 0.660 |

SD, standard deviation; Q1, first interquartile; Q3, third interquartile; Hb, hemoglobin; RBCs, red blood cells (total erythrocyte count); HCT, hematocrit; MCV, mean corpuscular volume; MCH, mean hemoglobin concentration; sTfR, soluble transferrin receptor; sTfR/log Ferr index, soluble transferrin receptor index/ferritin logarithm. The Wilcoxon signed rank test and paired Student's t test were performed.

Before treatment, more than half of the patients (n =22; 56.4%) had lower than normal serum iron levels; however, at the time of clinical cure, the percentage of patients with lower than normal serum iron levels was significantly reduced (**Table 4**). It was also noted that the mean values of these patients increased significantly with the use of antifungal treatment (**Table 5**). The sTfR levels and sTfR/log ferritin ratios before treatment and at the time of clinical cure were not significantly different (**Table 5**).

### Inflammatory process

More than half of the patients (n = 20; 62.5%) had values higher than the normal range before treatment (n = 32). At the time of clinical cure, the percentage of patients with values outside the normal range had not decreased significantly (n = 7; 21.9%; p = 0.225). However, an analysis of the median values of each disease stage showed a significant reduction in the values at the time of clinical cure compared to those before treatment (**Fig 3**).

**Fig 3.**
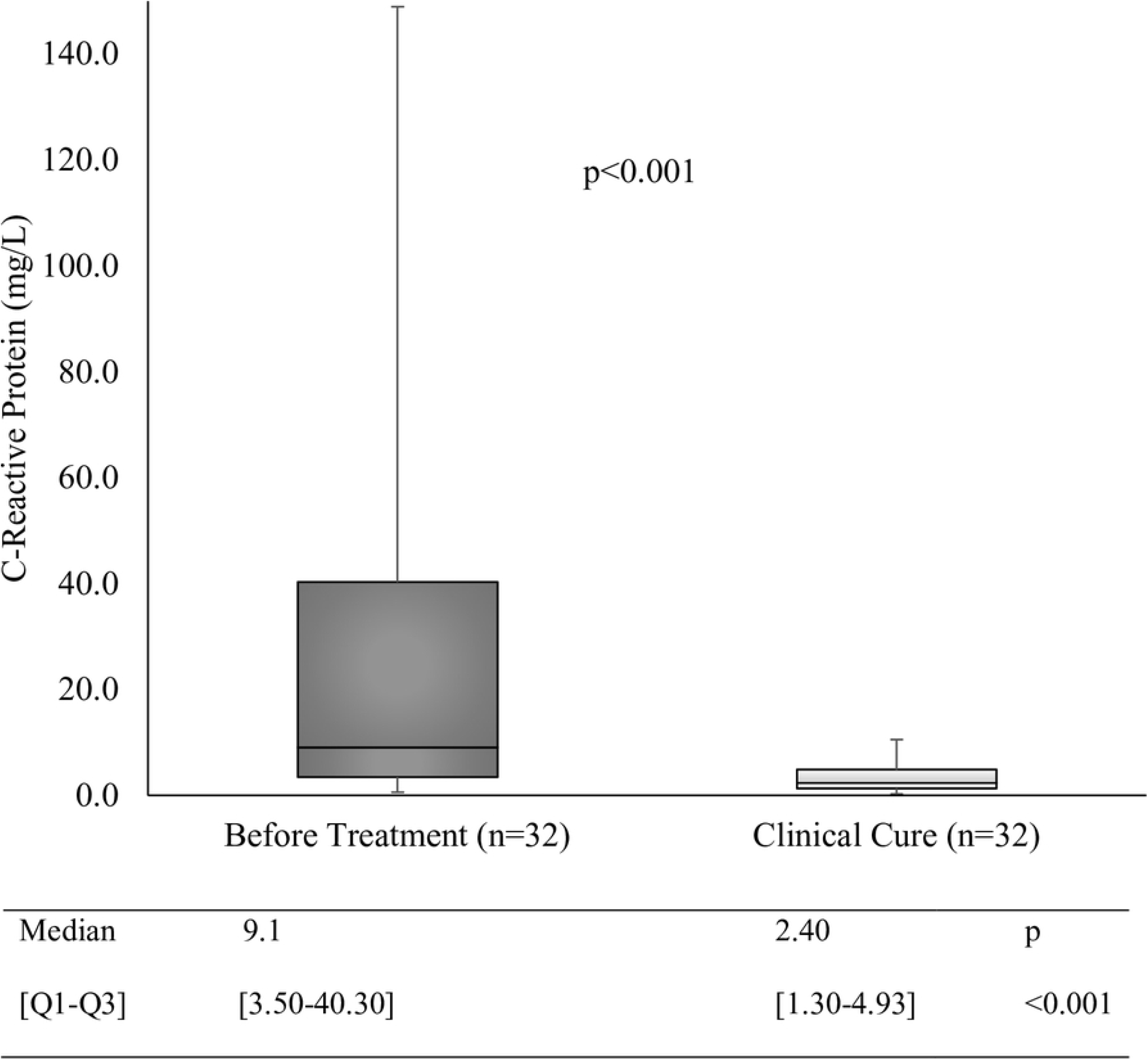
Boxplot of the inflammatory process evaluation involving CRP during different stages of PCM. Q1, first quartile; Q3, third quartile; BT, before treatment; CC, clinical cure. Wilcoxon signed rank test.

Weak to moderate indirect correlations between CRP levels during active disease (before treatment) and the TIBC (r = -0.405; p = 0.014), Hb level (r = -0.500; p = 0.002), RBC level (r = -0.461; p = 0.005), HCT level (r = -0.514; p = 0.001), and iron level (r = -0.491; p = 0.002) were observed (**Fig 4**). At the time of clinical cure, no correlations were observed between the CRP levels and these variables.

**Fig 4.**
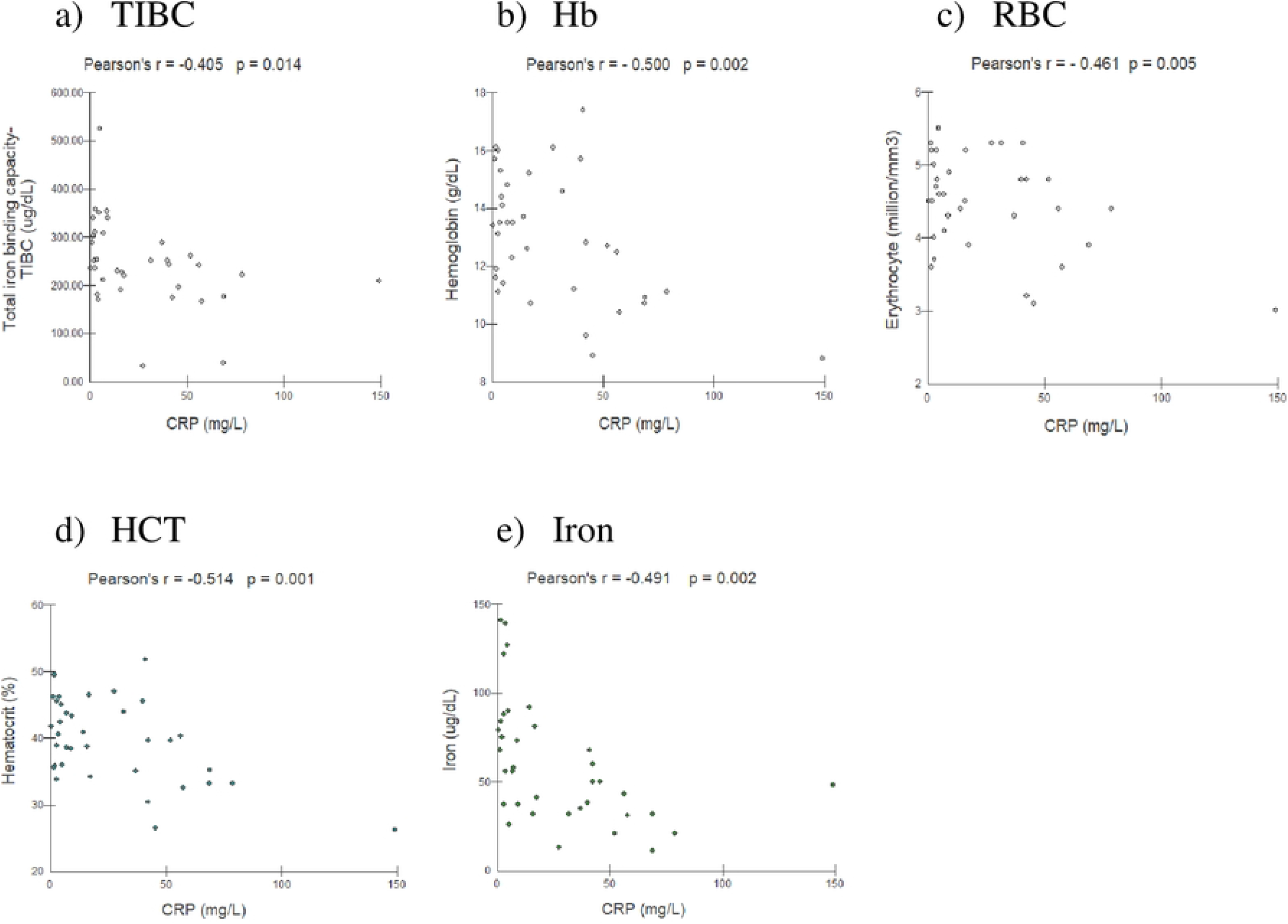
Pearson’s correlations of C-reactive protein (CRP) with variables used to evaluate the iron forms of patients with chronic paracoccidioidomycosis before treatment (n = 36 patients). (a) CRP with the total iron-binding capacity (TIBC). (b) CRP with hemoglobin (Hb). (c) CRP with red blood cells (RBCs; total erythrocyte count). (d) CRP with hematocrit (HCT).

An analysis of the stored iron and functional iron parameters before treatment showed a significant difference between values associated with severe and mild/moderate disease cases as well as CRP levels (**Table 6**).

**Table 6.** Analysis of iron parameters and CRP levels according to disease severity of patients with chronic paracoccidioidomycosis before treatment.

| Variables | Severe (n = 08) | Mild or Moderate (n = 31) | p |
| --- | --- | --- | --- |
|  | Mean (SD) or<br>Median [Q1-Q3] | Mean (SD) or<br>Median [Q1-Q3] |  |
| **Storage iron** |  |  |  |
| Ferritin | 448 [308-514] | 199 [90.4-290] | 0.007 |
| **Transport iron** |  |  |  |
| TSAT | 18 [14.3-22.8] | 27 [18-38.2] | 0.051 |
| TIBC | 215 [191-245] | 241 [201-310] | 0.258 |
| **Functional iron** |  |  |  |
| RBCs | 4.05 (0.81) | 4.60 (0.57) | 0.031 |
| Hb | 11.8 (3.1) | 13.5 (1.8) | 0.050 |
| HCT | 35.8 (8.8) | 41.0 (4.8) | 0.029 |
| Iron | 40.4 (14.5) | 68.1 (35.5) | 0.039 |
| **CRP** | 51.7 [40.8-71.4] | 7.06 [2.90-20.1] | <0.001 |

Q1, first quartile; Q3, third quartile; TSAT, transferrin saturation; TIBC, total iron-binding capacity; RBC, red blood cells (total erythrocyte count); Hb, hemoglobin; HCT, hematocrit; CRP, C-reactive protein. The Mann-Whitney U test and independent Student's t test were performed.

## Discussion

The acquisition of iron by microorganisms is an important virulence mechanism [16], and it has been reported with *Paracoccidioides* spp. [9, 17]. Additionally, human organisms actively control intracellular and systemic iron levels in a way that can contain infection and/or microbial persistence. Therefore, this study compared iron metabolism parameters before treatment and at the time of clinical cure, analyzed them based on PCM severity, and correlated them with the intensity of the inflammatory process in patients with chronic PCM. The demographic and clinical characteristics of the patients corresponded to those reported by other case series studies of chronic PCM that involved mostly male patients who were older than age 40 years, smokers, rural workers, and had moderate PCM [18-22]. Therefore, this sample is representative of the population of PCM patients and allows the generalization of our results.

The storage iron evaluated during this study had higher median serum ferritin levels before treatment, but they were within the normal range. Although ferritin exists in large amounts in the liver and spleen, only a small amount is detected in the circulation. However, serum quantification is a common and non-invasive method that provides an accurate measurement of storage iron [23]. However, the increase in serum ferritin may be related not only to increased storage iron but also to inflammatory activity. Because serum ferritin is an acute-phase protein in inflammatory and infectious processes and in cancer, the production of apoferritin is increased because of the stimulation of interleukins (ILs) such as IL-1 and IL-6 [24]. Increases in these ILs have been observed in patients with PCM [25].

During this study, serum ferritin levels were increased in some patients; however, the median values were within the normal range before treatment. At the time of clinical cure, these median values were significantly decreased, suggesting the interference of PCM with this parameter. Additionally, severe cases were associated with higher ferritin levels, probably because of the production of IL-6 caused by fungal infection. A previous study by our group reported similar results [14].

To assess the transport iron, we evaluated the TIBC, TSAT level, and serum iron/TIBC ratio. Before treatment, the patients in this study had median values that were slightly lower than normal, indicating a non-significant transport iron deficit. Additionally, antifungal therapy did not affect these parameters. A weak negative correlation was observed between the TIBC and CRP level, but no correlation was observed between the TSAT and CRP levels. The TIBC is an indirect measure of circulating transferrin, which can be reduced during inflammatory processes [11]. Therefore, with chronic PCM, the transport iron parameters are less affected than the storage iron and functional iron parameters.

A TSAT level <16% is considered indicative of iron deficiency with erythropoiesis; however, the use of this value for this parameter has limitations because both iron levels and the TIBC are lower with inflammatory processes [12, 23]. During our study, TSAT values <16% were found in 8 (20.5%) patients before treatment. However, only one of them had confirmed iron deficiency anemia, which is characterized by low Hb, ferritin, and TSAT levels [26]. A study of the effect of PCM treatment on iron metabolism showed that specific treatment quickly leads to a return to normal values or improvement in iron metabolism, as described with the TIBC, which was lower and normalized soon before antifungal treatment [27].

In clinical practice, the HMG test results and serum iron level are the first indicators of a change in the iron status. Although our study showed that the parameter values were slightly or not altered before treatment, it was possible to infer that PCM interferes with functional iron because these values improved after treatment and were associated with PCM severity. Additionally, a moderate correlation was observed between Hb, RBC, HCT, iron, and CRP levels. Furthermore, similar to our results, tuberculosis patients with active disease had lower mean values of these variables [28].

Approximately 65% of the total body iron in circulation in humans is linked to Hb [7], and reduced Hb with chronic infectious diseases is attributable to the limitation of erythropoiesis caused by regulators such as IL-1 and tumor necrosis factor-α [29]. Hb is a source of iron for *Paracoccidioides* spp., which removes iron by hemolytic activity and internalizes protoporphyrin rings through receptor-mediated pathways [30]. Other fungi are also capable of removing iron from Hb, such as *Candida albicans*, through a similar mechanism [31].

During infection, competition between the pathogen and host for iron uptake involves strategic mechanisms. Therefore, in response to infection, human organisms activate the immune system, thus inducing changes in circulating iron and leading to iron storage in macrophages and decreased absorption of iron from the diet by enterocytes [32]. Furthermore, several inflammatory cytokines are released by immune system cells, and they contribute to alterations in iron metabolism [33]. Despite the host-generated deprivation of iron, all fungi, including *Paracoccidioides* spp., have specific mechanisms for iron uptake through the production of molecules with high affinity for iron, siderophores, iron uptake pathways through reductive iron assimilation, or even Hb receptors [9, 34, 35]. Therefore, an inflammatory response associated with infection alters the availability of circulating iron, thus increasing its intracellular storage and leading to hypoferremia and iron-restricted erythropoiesis, as observed during our study, because these changes can contribute to anemia of inflammation [36].

The sTfR level is a good indicator of the iron status in the absence of systemic influences; therefore, we analyzed sTfR levels before treatment and at the time of clinical cure. Our results showed that the median values before treatment were normal and did not decrease after treatment, suggesting that the body iron is normal; however, it is distributed from functional iron to storage iron. A high sTfR level can indicate iron deficiency [37, 38]. During the storage iron reduction phase, the sTfR level does not change. However, when there is a decrease in functional iron, the synthesis of the transferrin receptor is stimulated; consequently, the sTfR level increases. This parameter is mainly recommended for differentiating iron deficiency anemia from inflammation anemia/chronic disease anemia because the value is increased with iron deficiency anemia but normal with inflammation anemia [39-41]. Studies of pulmonary tuberculosis patients suggested that high sTfR levels in patients with anemia indicate probable iron deficiency and recommended that the sTfR concentration of patients with anemia and an infectious/inflammatory process should be examined [28].

Another good indicator of iron deficiency with differential anemia is the sTfR/log ferritin ratio [42]. During our study, the sTfR/log ferritin ratio as well as sTfR level were normal before treatment and did not significantly change after treatment, reinforcing the belief that anemia with chronic PCM is inflammation anemia.

This study presents some limitations. The number of patients is apparently low. However, our Service receives about 15 new cases every year and most of the patients do not undergo the follow-up after clinical improvement, becoming difficult the pairing of some variables – before treatment and after clinical cure.

In conclusion, PCM interferes with iron metabolism by shifting functional iron to storage iron, as revealed by anemia and low iron levels, with normal TSAT and sTfR levels, normal TIBC, and normal sTfR/log ferritin ratio. Normal or slightly increased ferritin levels, which are characteristic of inflammatory anemia, were observed. However, it should be noted that a few patients had iron deficiency anemia. Therefore, a careful investigation of the iron parameters is important to the clinical management of PCM.

## Data Availability

All relevant data are within the manuscript and its Supporting Information files.

## Supporting information

**S1 Dataset. Dataset containing variables of the study**.

## Notes

### Competing Interest Statement

The authors have declared no competing interest.

### Funding Statement

ECAB and AOGMS: Coordenação de Aperfeiçoamento de Pessoal de Nível Superior – Brasil (CAPES) – Código de Financiamento 001. AMMP: Ministério da Ciência, Tecnologia e Inovação and Conselho Nacional de Desenvolvimento Científico e Tecnológico (grant numbers: 312910/2020–7 and 431,776/2016–4) and Fundação de Apoio ao Desenvolvimento do Ensino, Ciência e Tecnologia do Estado de Mato Grosso do Sul (grant number: 71/000.478/2021). IVS: Federal University of Mato Grosso do Sul VTLF: Conselho Nacional de Desenvolvimento Científico e Tecnológico

### Author Declarations

Human Research Ethics Committee of the Federal University of Mato Grosso do Sul (CAAE number 62726016.2.0000.0021)

